# Analysis of State-level Variation in Distribution of Oxycodone and its Adverse Effect Profile in the US from 2000-2021

**DOI:** 10.1101/2022.09.03.22278873

**Authors:** Jay P. Solgama, Edward Y. Liu, Mellar P. Davis, Jove Graham, Kenneth L. McCall, Brian J. Piper

## Abstract

**Objectives:** Characterize oxycodone’s distribution in the US by state and its adverse effect profile from 2000-2021.

**Design:** Observational

**Setting:** More than 80,000 Americans died of an opioid overdose in 2021 as the United States (US) continues to struggle with an opioid crisis. Prescription opioids play a substantial role, introducing patients to opioids and providing a supply of drugs that can be redirected to those seeking to misuse them.

**Methods:** The Drug Enforcement Administration annual summary reports from the Automation of Reports and Consolidated Orders System (ARCOS) provided weights of oxycodone distributed per state by business type (pharmacies, hospitals, and practitioners). Weights were converted to Morphine Milligram Equivalents (MME) per capita and normalized for population.

**Results:** There was a sharp 280.13% increase in total MME/person of oxycodone from 2000-2010, followed by a slower 54.34% decrease from 2010-2021. Florida (2007-11), Delaware (2003-20), and Tennessee (2012-21) displayed consistent and substantial elevations in combined MME/person compared to other states. In the peak year (2010), there was a 15-fold difference between the highest and lowest states. MME/person from only pharmacies, which constituted >94% of the total, showed similar results. Hospitals in Alaska (2000-01, 2008, 2010-21), Colorado (2008-21), and DC (2000-11) distributed substantially more MME/person over many years compared to other states. Florida stood out in practitioner-distributed oxycodone, with an elevation of almost 15-fold the average state from 2006-2010. The percentage of consumer reports in the FDA Adverse Drugs Events Reporting System (FAERS) increased from 6.3% (2001) to 95% (2021). Adverse effects distribution remained constant by age and sex, with higher average proportions in males (55%) and the 18-65 age group (79%).

**Conclusions:** Oxycodone distribution across the US showed marked differences between states and business types over time. Investigation of opioid policies in states of interest may provide insight for future actions to mitigate opioid misuse.

**Strengths and limitations of this study:**

- ARCOS is publicly accessible, comprehensive, and includes institutions that are unavailable in other commonly used databases (i.e. IQVIA).
- Diversion of oxycodone or whether the prescribed amounts were utilized cannot be determined from ARCOS data.
- FAERS data includes duplicates, incomplete results, and non-verifiable data which cannot be used to suggest causation between oxycodone and adverse events.
- These complementary results from two databases may not generalize to other countries with more restrictive oxycodone policies.

## INTRODUCTION

The United States (US) is amid an opioid crisis that has been worsening since the start of the century. The most recent data shows that 80,725 individuals died from an opioid overdose in 2021.^1^ Prescription opioids put pain management patients at risk for developing an addiction and provide a potential source for opioid misusers. Since the 1990s, many people were introduced to opioids through prescription drugs, and some moved on to cheaper alternatives such as heroin.^2^ Regulations and guidelines often apply at the state level, and heterogeneity in culture, politics, and socioeconomics play a role in the disproportionate impact of the crisis in different regions. In this study, we aimed to characterize the US distribution of oxycodone, a prevalent opioid that has contributed to the crisis, across states.

Oxycodone was the most commonly consumed opioid globally from 2009-19, with the US being the largest consumer.^3^ The generic product and other formulations (e.g., OxyContin, Percocet, Percodan, Roxicodone) were misused by 1.1% of the US population in 2020, behind only hydrocodone.^4^ Oxycodone is of particular concern due to its high abuse liability.^5^ Its danger is demonstrated by high rates of overdose death, as it was involved in 33,154 deaths between 2011-16 and is the third highest on the list behind heroin and fentanyl.^6^

Few studies have investigated trends in oxycodone distribution throughout the US. One investigation utilized the Drug Enforcement Administration’s Automation of Reports and Consolidated Orders System (ARCOS) database, finding a concerning 69.7% increase in amount of oxycodone distributed from 2000-2010.^7^ A more recent report performed a detailed analysis for both oxycodone and hydrocodone in US commuting zones using Washington Post ARCOS data found that population size influenced scaling behavior of pills, and discovered regions of high sales in the Appalachians, Ozarks and the west coast.^8^

Here, we explore oxycodone distribution by state using the US Drug Enforcement Agency (DEA) ARCOS over a larger time frame than any previous studies, which included reports on prescription opioids in the US territories, Texas, and throughout Delaware, Maryland, and Virginia.^9–11^ Past works frequently rely on the IQVIA database, which is limited by gaps in oxycodone, including data from Veteran’s Affairs, Indian Health Services facilities, hospitals, and independent pharmacies.^12^ Using states as the geographical unit allows results to be interpreted in the context of laws and regulations which apply at that level. We include analysis of distribution by three business types: pharmacies, hospitals^13^, and practitioners, to provide more detailed information. We also utilize the FDA Adverse Drugs Events Reporting System (FAERS), an information database that contains adverse drug events (ADEs), medication error reports, and product quality complaints submitted to FDA from healthcare professionals, consumers, and others (i.e. manufacturers) through MedWatch.^14^ We report on the many ADEs, including dry mouth, constipation, nausea, and vomiting to severe ones like hallucinations, agitation, chest pain, allergic reactions, and death for oxycodone.^15^

## METHODS

### Procedures

Data was collected from the ARCOS Report 5. The database is provided by the DEA and reports on distribution of Schedule II and III controlled substances.^16^ Oxycodone distribution was reported in grams grouped by business type (hospitals, pharmacies, practitioners, mid-level practitioners, narcotic treatment programs, teaching institutions) per state for 2000-2021 alongside the number of buyers. Oxycodone from teaching institutions was addressed elsewhere.^17^ Pharmacies, hospitals, and practitioners were chosen for individual analysis in addition to the total of all business types. The FDA Adverse Drugs Events Reporting System (FAERS) is a database containing adverse events reports that are submitted to the FDA. It was queried to analyze oxycodone through the search terms “oxycodone” and “oxycodone hydrochloride”; these terms include both mono and combo forms of the opioid (i.e. oxycodone, oxycodone and acetaminophen) in the database. Hereafter, “oxycodone” refers to both mono and combo products. This approach was used to analyze oxycodone adverse events from 2000 to 2021 to examine the proportion of ADEs with respect to demographics (sex, reporter type, and age group). We excluded the “unspecified” counts for each group which might consist of unverified or incomplete reports (Supplementary Table 1). The percentage of the top five reaction groups for both mono and combo oxycodone were analyzed to compare the distribution pattern of ADEs for both products.

### Data analysis

Microsoft Excel, GraphPad Prism, and RStudio were used to analyze and visualize the data. Grams of oxycodone were converted to morphine milligram equivalents (MME) using a standard oral conversion factor of 1.5.^18^ The total MME per state was divided by the corresponding year’s population, as reported by the U.S. Census Bureau’s American Community Survey, to calculate MME per person. MME per buyer was calculated by dividing MME and the provided number of businesses that made purchases.

MME per person was plotted across time for each state to identify the peak year of oxycodone distribution. The five states with the highest average distribution and the five states with the lowest average distribution were plotted for pharmacies, hospitals, and the total of all business types. Data for practitioners was much more varied, and ten states with the highest distribution and notable patterns were plotted.

States whose values that fell outside the range of ± 1.96 standard deviations from the mean were identified, and a Pearson correlation coefficient, r, was calculated for MME per person and MME per buyer for each business type (Supplementary Figure 1).

## RESULTS

### Total Oxycodone Distribution

The United States distributed 1,404.01 metric tons in MME of oxycodone from 2000 to 2021. Total MME per person increased rapidly and by +280% from 2000 to the peak in 2010. There was a more protracted decrease of -54% from 2010 to 2021. Notable differences arose when distribution was grouped by business type. Pharmacies constituted the largest proportion of oxycodone distribution (>94%) and drove the pattern of total distribution. Pharmacies showed a +293% increase from 2000-10 and a -54% decrease from the peak to 2021. Hospitals were far less variable in their distribution between years, with an increase of +80% to a later peak in 2012 and a subsequent slow decline of -51% by 2021. Practitioners showed a pronounced increase in their oxycodone distribution by +15,567% leading up to 2010 but declined by -87% the following year. There was a -99% reduction from the peak by 2021. MME per person was positively and strongly correlated with MME per buyer for each grouping, with an r value of +0.95 for all businesses, +0.9383 for pharmacies, +0.70 for hospitals, and +0.85 for practitioners.

### Oxycodone Distribution by State

MME per person distributed by states showed large variations between each other, in total, and for each business type. The difference between population corrected distribution was 8-fold in 2000 (Alaska = 177.7, Illinois = 23.4), peaked at 15-fold in 2010 (Florida = 993.5, Texas = 66.7), and decreased to 6-fold in 2021 (Tennessee = 250.8, Illinois = 45.2). Florida (2007-11), Delaware (2003-20), and Tennessee (2012-21) showed consistent and substantial elevations in combined MME/person compared to other states (i.e., greater than 1.96 standard deviations higher than the mean). Texas (2012-21) and Illinois (2006, 2013-21) were on the other end of the spectrum, with considerably lower oxycodone distribution.

MME per person from only pharmacies constituted the preponderance (>94%) of the total and displayed a similar pattern as the combined distribution. Florida (2007-11), Delaware (2003-20), and Tennessee (2012-21) had substantially higher values, while Texas (2012-21) and Illinois (2006, 2013-21) had substantially lower values, over many years. Most states showed some growth leading up to the years around the national peak in 2010 followed by a decline until 2021.

States tended to peak in the few years following 2010 when examining oxycodone distributed by hospitals. There were still large differences in magnitude between states. Hospitals in Alaska (2000-01, 2008, 2010-21), Colorado (2008-2021), and DC (2000-11) purchased the largest quantities of oxycodone. Only Illinois distributed amounts of oxycodone that were lower than 1.96 standard deviations from the mean, from 2000-02.

The data for practitioners showed notable variations between states in the magnitude of oxycodone distribution as well as patterns of change. Florida stood out with nearly 15-fold the MME/person for the average state from 2006-10. Florida was also 26,152-fold elevated relative to West Virginia in 2010. Delaware showed a sharp spike in 2011 that dropped back within ±1.96 standard deviations of the mean by 2013. Hawaii had notably high values in more recent years from 2014-15 and 2017-21. No states distributed amounts of oxycodone that were 1.96 standard deviations lower than the mean.

### Oxycodone Adverse Events

The distribution of adverse effects by sex remained relatively constant in a 1:1 ratio with the highest proportion of males in 2002 (63%) and females in 2017 (54%). The average percentage of males and females within the time interval was 55% and 45% respectively. When examined by age, the distribution of ADEs in individuals from 0-17 years old on average was infrequent (4%), >65 were higher (17%), and 18-64 accounting for the majority (79%).

There was a general increase in consumer reports (+1,407.9%) from low (6.3%) in 2001 and peak in 2021 (95.0%). Healthcare professional reports decreased over time in a reciprocal manner. The average percentage of consumer reports and healthcare provider reports within the time interval was 40.4% and 59.6% respectively.

The top five disorders for oxycodone from 2000 to 2001 were psychiatric disorders (26%), general disorders and administration site conditions–such as hypersensitivity reactions, fatigue, or malaise–(21%), injury, poisoning, and procedural complications (20%), nervous system (5%), and gastrointestinal disorders (4%), making up the top five reaction groups (>75%) and <25% for other reaction groups (Supplementary Table 2).

## DISCUSSION

This study utilized the comprehensive ARCOS database to characterize the US distribution of oxycodone by pharmacies, hospitals, practitioners, and in total for two decades. We saw a large increase in total oxycodone distribution from 2000 until 2010, during which time MME per person grew by four-fold. This was followed by a decrease by more than half over the next eleven years. There were stark differences in the patterns of change between states. Some, like Florida^11^ or Delaware^10^, increased substantially leading up to 2010 and dropped off just as quickly. Others like Texas^9^ or Illinois maintained consistently low distribution, and many states fell somewhere in between. Differences between states peaked at nearly fifteen-fold in the peak year (2010), and never dropped below five-fold. It is important to put the peak oxycodone MME in Delaware (918 MME/person) in context. The MME for ten opioids including oxycodone in 2016 in North Dakota was almost half (485) of this.^18^ These pronounced and persistent state-level disparities may offer opportunities for continued vigilance in opioid stewardship.

Pharmacies distributed nearly 95% of the oxycodone over the timeframe of the data, following a pattern close to the total distribution. Again, many states displayed large increases leading up to the peak year, preceding a slow decline. Comparison of the magnitude of distribution revealed about seven-fold, fifteen-fold, and six-fold differences in 2000, 2010, and 2021 respectively. Practitioner distribution also showed pronounced variation between states. Differences started at 928-fold in 2000, peaked at an astounding 26,152-fold in 2010, and settled at 3,192-fold in 2021. Overall, there was a rapid increase and decrease around the peak, but many states showed multiple peaks while others (e.g., West Virginia, Rhode Island, Montana) distributed relatively low amounts of oxycodone for all years. Oxycodone distributed by hospitals^13^ showed a far less drastic peak in 2012, later than the other groups, and slowly declined until 2021. Interestingly, differences between states peaked at 20-fold much earlier in 2005, eventually decreasing to 10-fold in 2021.

The distribution of oxycodone peaking in 2010 is possibly due to a reformulation of OxyContin to a more abuse-deterrent form which was harder to crush, for nasal insufflation, and dissolve, for intravenous injection.^19^ There is support for this change decreasing the misuse potential of the drug^20,21^, likely leading to decreased demand for misusers. Also in 2010, there was legal action taken against “pill mill” physicians, who inappropriately prescribed controlled medications, particularly in Florida.^22^ In the following years, attempts to decrease these harmful practices extended to many other states. However, it is also important to recognize that prior ACROS analyses noted that the peak was only slightly later (2011) for ten Schedule II opioids^18^ indicating that provider, payer, and patient attitudes towards prescription opioids have undergone substantial changes over the past decade.^23^

Oxycodone is mainly administered orally but can be given intravenously and rectally. Its uses as an immediate release formulation and IV injection can lead to higher misuse, and therefore greater chance of general and administration site problems (hypersensitivity and systemic reactions), injury, and poisoning^24^ The high proportion of psychiatric disorders may be explained by its predilection for the brain with previous studies indicating that is more concentrated in the brain than morphine, and greater relative amounts in the blood.^25^ The increase in consumer reports relative to healthcare professionals submissions over time, peaking in 2021 (95%) is surprising. Consumers may have had more time compared to healthcare professionals to submit reports and may report more frequently due to quality-of-life related adverse events given oxycodone’s high potential for misuse.^26^ Furthermore, young to middle-aged adults have higher rates of opioid-related adverse effects, specifically 1 in 5 deaths of the 25-34 age group due to opioids in the US.^27^ The large proportion of individuals within that age group may also contribute to the relatively large percentage of ADEs for the 18-64 age range.

The US stands out considerably from other countries in the world regarding oxycodone consumption. In 2020, the US accounted for 68.2% of global oxycodone consumption, dwarfing the other major consumers, Germany (5.2%), Canada (3.2%), France (3.1%), China (2.8%), Australia (2.4%), and the United Kingdom (2%).^28^ Regulations in the US also differ greatly from other countries, where national regulations are more common and guidelines have a larger impact on clinical practice.^29^ Although the US develops guidelines at a national level, individual states are responsible for their implementation. A 2019 report from the Organization for Economic Cooperation and Development (OECD) took an in-depth look at the opioid crisis and policies to address it in its 38 member countries.^30^ They found that several evidence-based policies were effective in improving outcomes in some countries or US states, such as patient education programs, training/education initiatives for providers, interventions against stigma, naloxone programs, and syringe programs. Opioid prescribing guidelines in the US have been shown to be effective in mitigating opioid prescriptions.^31^ Although there is much research to be done to validate and develop methods to reduce harm from opioid misuse, there are already actions that can be taken to save lives.^32^

There are some strengths and limitations to this novel report due to the nature of the data. The ARCOS data is accessible by the public, and provides pharmacoepidemiological information that includes Veteran’s Affairs, Indian Health Services facilities, hospitals, and independent pharmacies that are unavailable in the IQVIA database.^12^ However, the database includes all transactions and may be a slight overestimate due to shipments between the same distributors.^33^ A modest amount of oxycodone reported by ARCOS in the pharmacy, practitioners, and teaching institutions^17^ business activity is used by veterinarians. The database provides licit distribution of oxycodone, and any diversion of the drug to unintended recipients is not publicly available. Similarly, it cannot be determined if patients utilized the quantity of drug that they were subsequently prescribed. The American Community Survey data is a likely underestimate of actual population due to limitations in counting undocumented individuals.^34^ Although oxycodone was among the most prescribed and misused opioids from 2000-21 (12,289,518 prescriptions in 2020)^35^, further research with greater spatial resolution within a single state (e.g., Florida) or on other opioids may provide valuable information.

The adverse events from the FAERS database do not suggest causation between that of the drugs and the event. Both mandatory (manufacturers of drugs) and voluntary (i.e. consumers and healthcare professionals) report adverse event reports to the system, but the FDA cautions its use due to a few reasons. The reports provide descriptive data rather than definitive and might not be generalizable.^14^ Reports may also be overrepresented because of duplicates, incomplete results, or non-verifiable data.^14,36^ The side effect profiles for oxycodone products may vary between consumers and healthcare professionals due to factors like the completeness of reports, types of adverse events reported, and frequency of reports.^26^ Though unspecified reports were excluded from our study, they made up a large percentage in the age group.

Future work can build upon these novel findings by interpreting them through the lens of guidelines and regulations that control the prescribing and distribution of oxycodone as well as justifiable hesitation among some patients to be prescribed opioids for chronic pain. The results of this report can provide beneficial information for development and revision of public health policies. Guidance from governmental agencies is necessary to address problems that are as pervasive and widespread as the opioid epidemic. This oversight is constantly evolving including with the implementation of the CDC’s 2022 update of its Clinical Practice Guideline for Prescribing Opioids.^37^ It is also important to view this data in relation to the escalating problem that needs to be more fully addressed, opioid overdose deaths.^1^ The process of decreasing availability of prescription opioids holds the risk of increasing demand from illicit sources, potentially increasing deaths. Investigating the socioeconomic and geopolitical context of states during changes in oxycodone distribution and overdoses may shed some light on how to approach the opioid epidemic in the coming years as the US continues to correct for prior excesses.^38,39^ We are cautiously optimistic that other countries will continue to be more judicious with oxycodone than the US.

## Data Availability

ARCOS data is publicly accessible at https://www.deadiversion.usdoj.gov/arcos/retail_drug_summary/.

FAERS data is publicly accessible at https://www.fda.gov/drugs/questions-and-answers-fdas-adverse-event-reporting-system-faers/fda-adverse-event-reporting-system-faers-public-dashboard.

A python script to extract data from ARCOS Report 5 PDFs (provided at: https://www.deadiversion.usdoj.gov/arcos/retail_drug_summary/index.html) and process the data can be found at https://github.com/solgamaj/VARCOS

## Acknowledgements

Audrey L. Valentine is appreciated for her contribution to data collection.

## Author Statement

Jay P. Solgama: Conception and design of the study, acquisition and analysis of ARCOS data, drafting of the manuscript.

Edward Liu: Acquisition and analysis of FAERS data, drafting of relevant sections.

Mellar P. Davis, MD: Revision and approval of the manuscript.

Jove Graham, PhD: Revision and approval of the manuscript.

Kenneth L. McCall, PharmD: Revision and approval of the manuscript.

Brian J. Piper, PhD, MS: Conception and design of the study, revision and approval of the manuscript.

## Data Sharing Statement

ARCOS data is publicly accessible at https://www.deadiversion.usdoj.gov/arcos/retail_drug_summary/.

---

**Figure 1.**
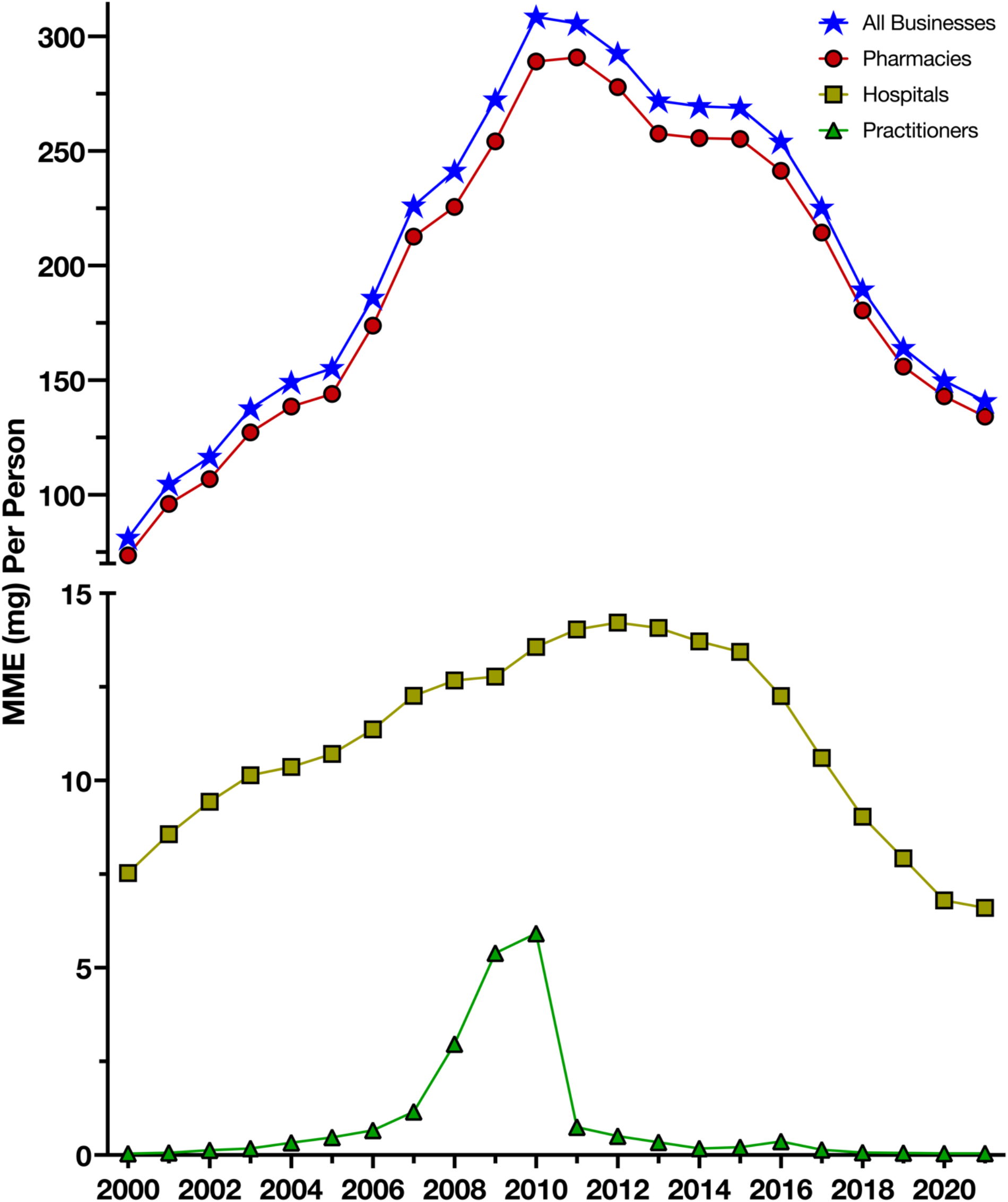
Total oxycodone distribution and by business activity in the US from 2000 to 2021 as reported by the Drug Enforcement Administration’s Automated Reports and Consolidated Orders System.

**Figure 2.**
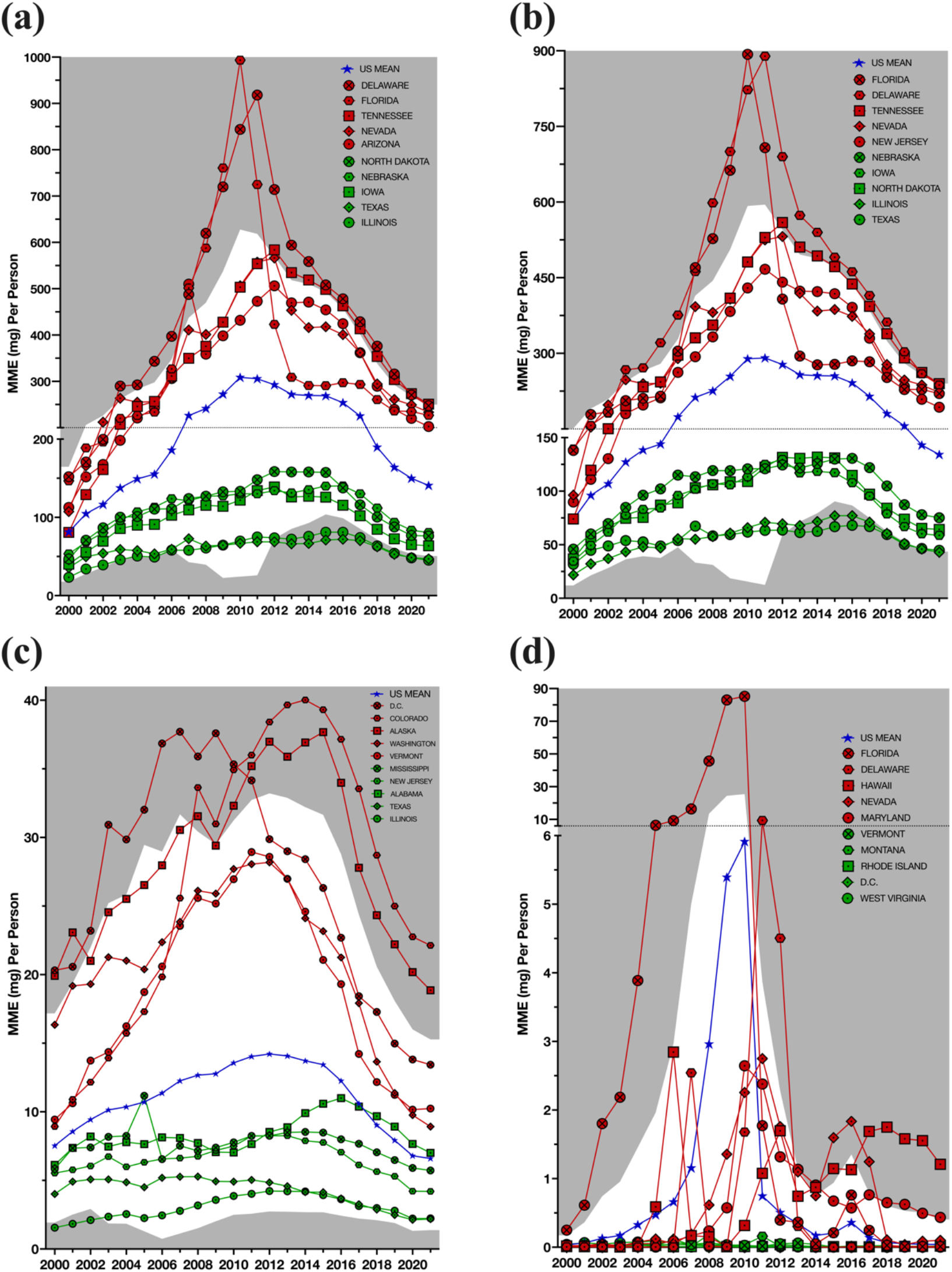
Morphine mg equivalent (MME) per person of oxycodone by business type 2000-2021 as reported by the United States Drug Enforcement Administration’s Automated Reports and Consolidated Orders System for (a) All Business Types, (b) Pharmacies, (c) Hospitals, and (d) Practitioners. Areas outside ±1.96 standard deviations of the average state are in gray.

**Figure 3.**
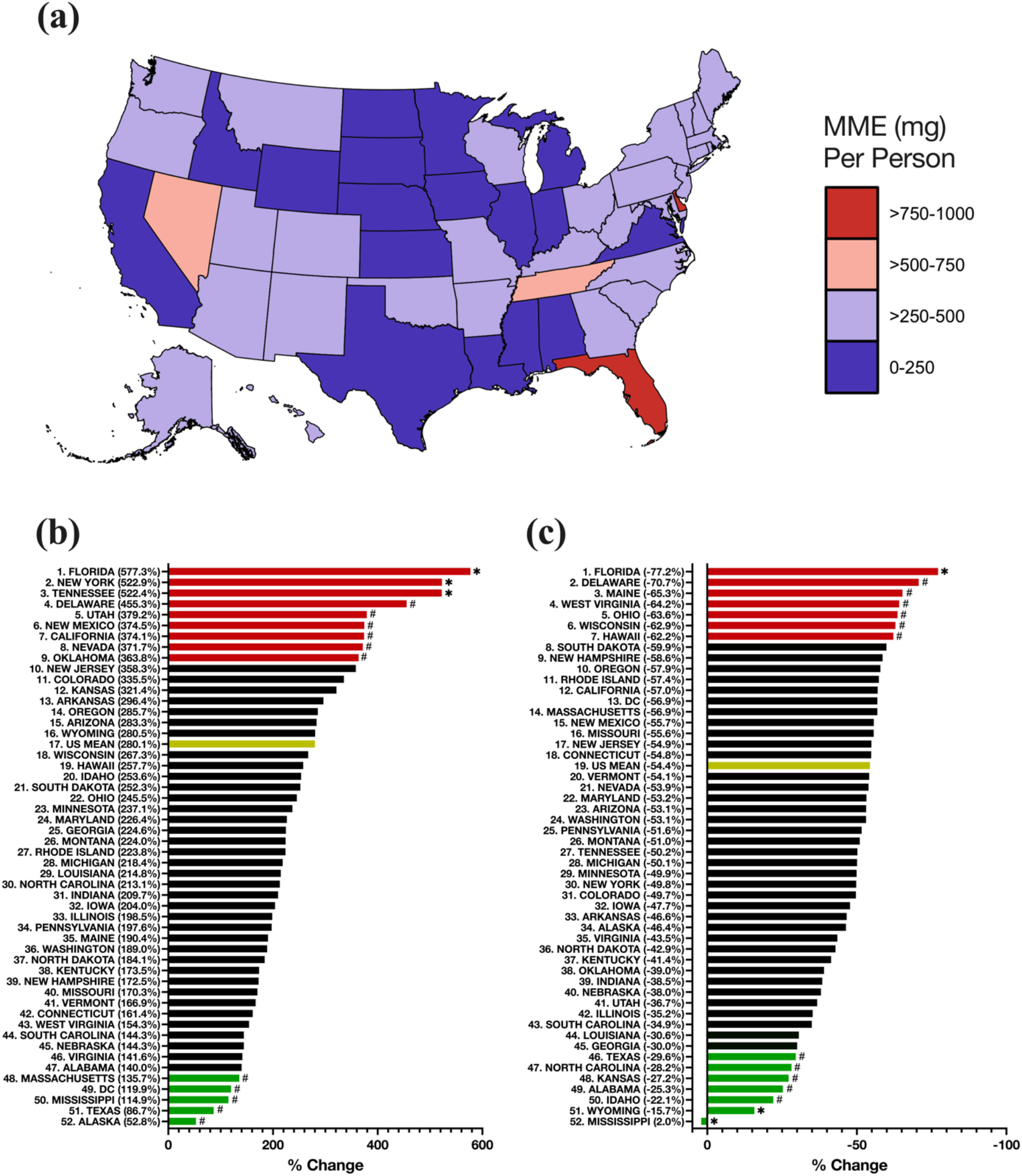
Heat map of the morphine mg equivalent (MME) per person in the peak year (2010) as reported by the Drug Enforcement Administration’s Automated Reports and Consolidated Orders System (a). Bar graphs showing the percent increase from 2000 to 2010 (b) and percent decrease from 2010-21 (c). States outside ^*^±1.96 or ^#^±1.0 standard deviations of the mean.

**Figure 4.**
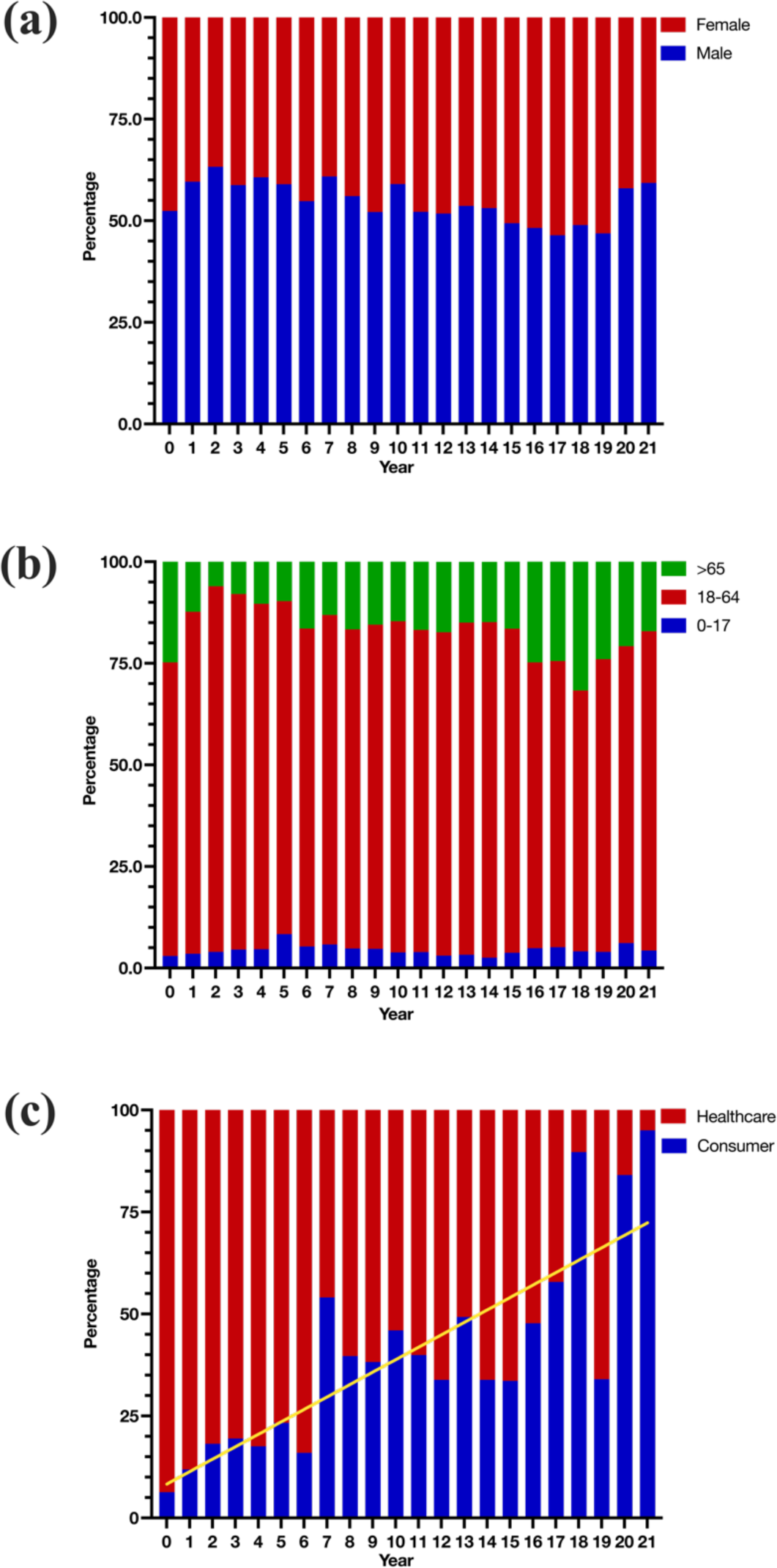
Percentage by demographics (sex (a), age group (b), and reporter type (c)) of oxycodone adverse drugs events (ADEs) in the US from 2000-2021 as reported by the FDA Adverse Drugs Events Reporting System (FAERS). “Healthcare” refers to healthcare professional reports.

**Figure 5.**
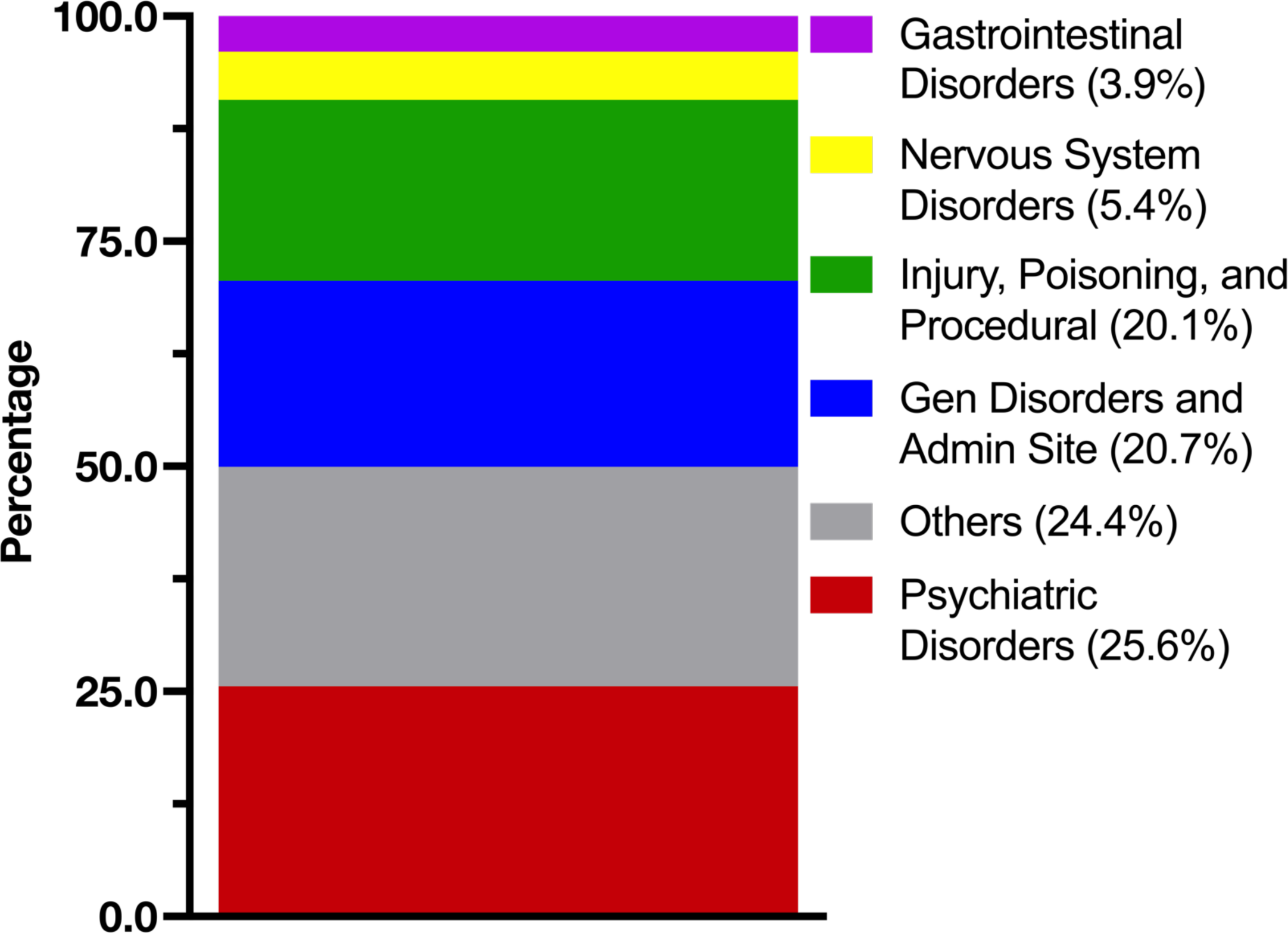
Percentage of oxycodone adverse drug events as reported by the FDA Adverse Drugs Events Reporting System for the top five categories. Other included cardiac, respiratory, thoracic, and mediastinal disorders.

