## Supplemental Figure 1 for "Analysis of State-level Variation in Distribution of Oxycodone and its Adverse Effect Profile in the US from 2000-2021"

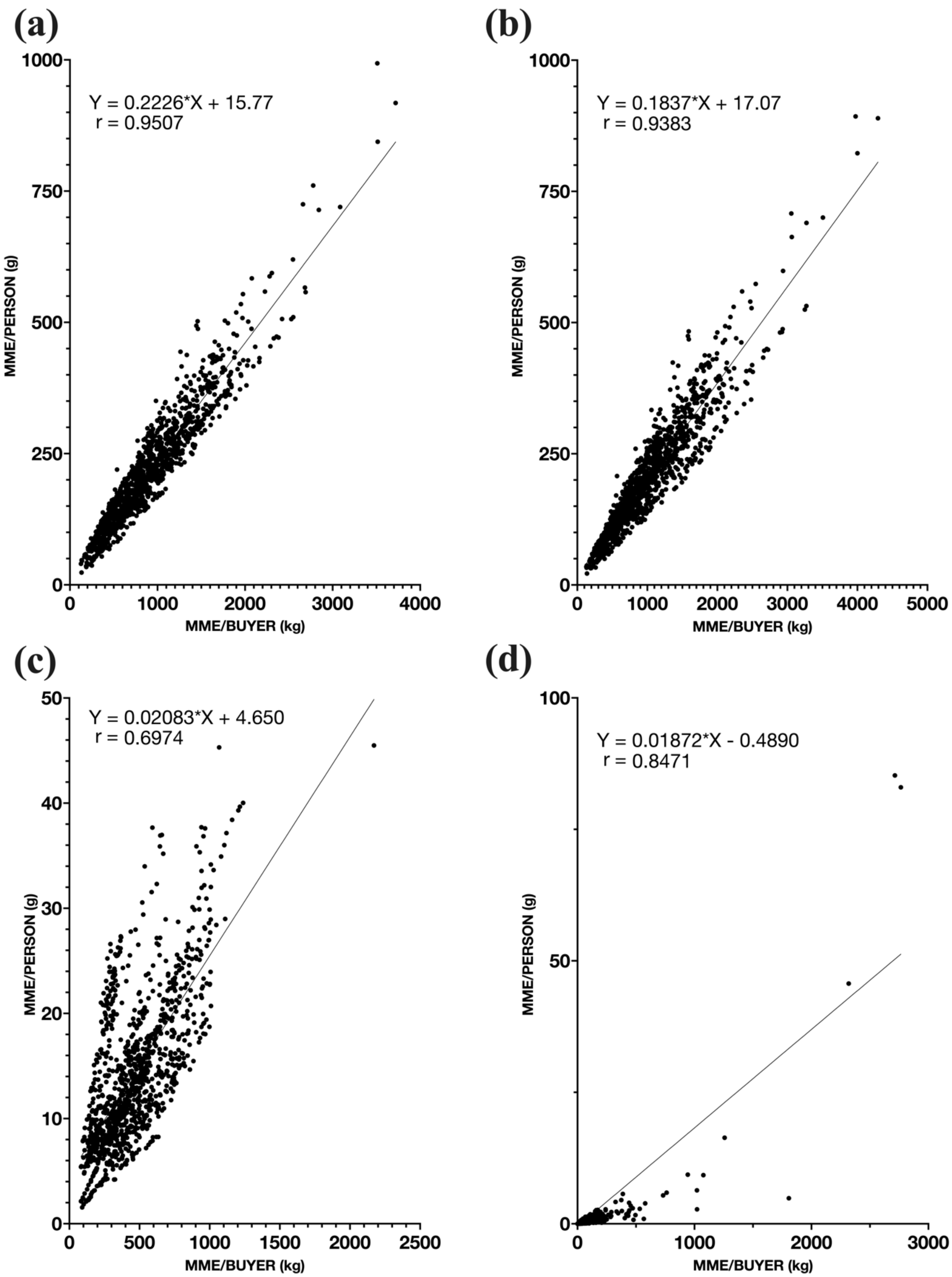


**Supplementary Figure 1**. Correlations between the morphine mg equivalent (MME) per person and the MME per buyer for (a) all businesses. (b) pharmacies. (c) hospitals, and (d) practitioners.
