## Supplemental Figure 2 for "Analysis of State-level Variation in Distribution of Oxycodone and its Adverse Effect Profile in the US from 2000-2021"

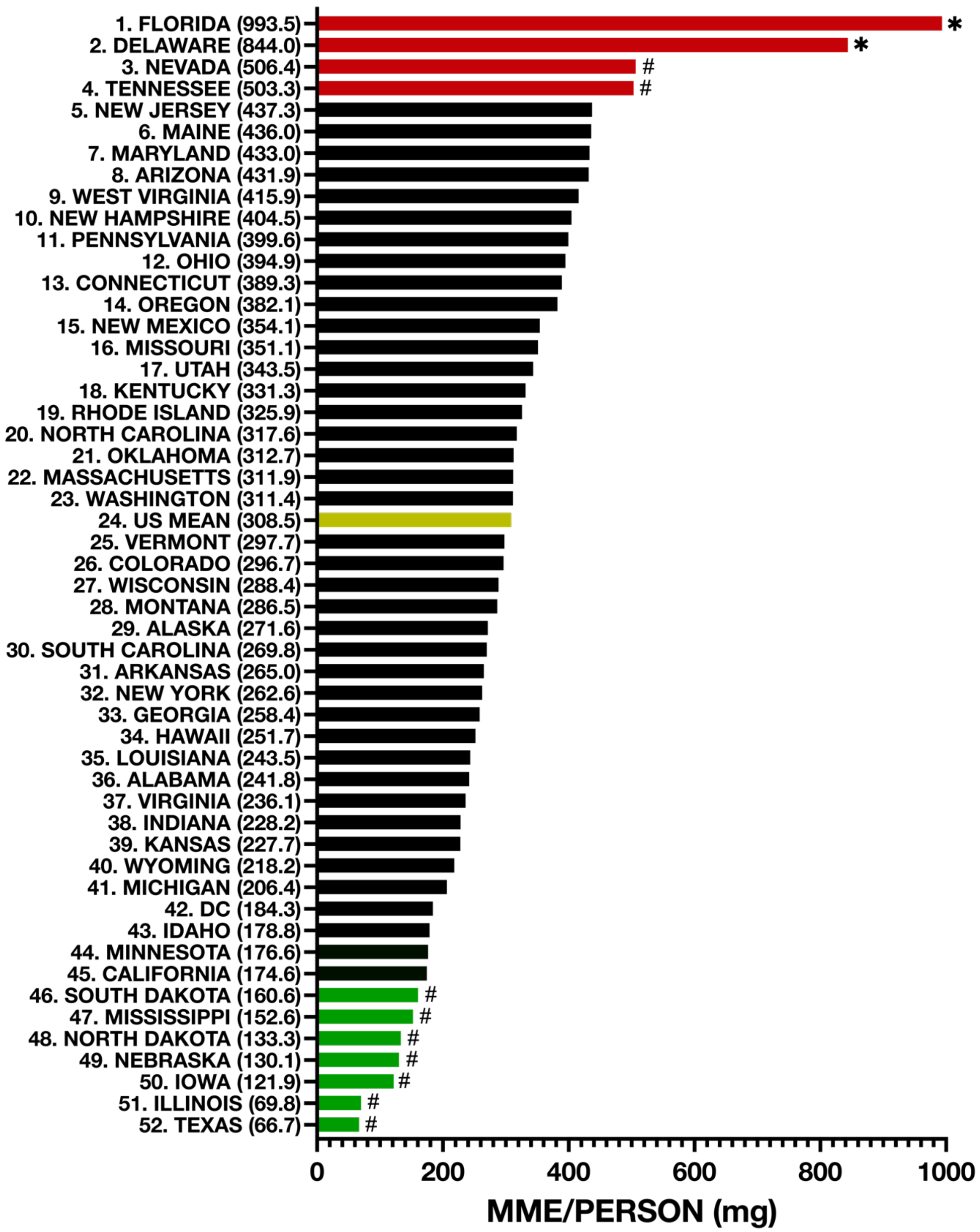


**Supplementary Figure 2.** Bar graph of the morphine milligram equivalent (MME) per person in the peak year 2010 with rank and MME/person in milligrams for each state. States outside $^{*}\underline{+}1.96$ or $^{\#}\underline{+}1.0$ standard deviations from the average state.
