## Supplemental Table 1 for "Analysis of State-level Variation in Distribution of Oxycodone and its Adverse Effect Profile in the US from 2000-2021"

**Supplemental Table 1.** Total number of cases of adverse drug events (ADEs) in the US from 2000-2021 as reported by the FDA Adverse Drugs Events Reporting System (FAERS) of sex, reporter type, and age group after excluding unspecified counts for each demographic group.

| **Year** | **Total cases** | **Sex #** | **Reporter #** | **Age Group #** |
| --- | --- | --- | --- | --- |
| 2000 | 109 | 105 | 95 | 101 |
| 2001 | 680 | 651 | 596 | 569 |
| 2002 | 1763 | 1707 | 1425 | 1431 |
| 2003 | 3581 | 3342 | 3273 | 2336 |
| 2004 | 3941 | 3656 | 3479 | 2172 |
| 2005 | 3061 | 2834 | 2655 | 2072 |
| 2006 | 1237 | 1095 | 957 | 938 |
| 2007 | 2083 | 1986 | 1415 | 1100 |
| 2008 | 1319 | 1197 | 1199 | 997 |
| 2009 | 1246 | 1137 | 1154 | 912 |
| 2010 | 2073 | 1987 | 1896 | 1510 |
| 2011 | 1895 | 1786 | 1831 | 1455 |
| 2012 | 1961 | 1844 | 1911 | 1505 |
| 2013 | 7706 | 7526 | 7619 | 3655 |
| 2014 | 4869 | 4740 | 4753 | 3270 |
| 2015 | 4160 | 3919 | 4121 | 2822 |
| 2016 | 3872 | 3625 | 3852 | 2233 |
| 2017 | 6148 | 4108 | 6130 | 2554 |
| 2018 | 27462 | 4549 | 27430 | 2993 |
| 2019 | 6420 | 5668 | 6386 | 4093 |
| 2020 | 24400 | 22125 | 24372 | 6072 |
| 2021 | 62012 | 59660 | 61977 | 5008 |
