## Supplemental Table 2 for "Analysis of State-level Variation in Distribution of Oxycodone and its Adverse Effect Profile in the US from 2000-2021"

**Supplemental Table 2.** Percentage breakdown of reaction groups of the “Other” category for oxycodone as reported by the FDA Adverse Event Reporting System.

| **Reaction Group** | **Percentage** |
| --- | --- |
| social circumstances | 2.919 |
| respiratory, thoracic, and mediastinal disorders | 2.730 |
| cardiac disorders | 2.601 |
| investigations | 2.317 |
| musculoskeletal and connective tissue disorders | 1.724 |
| infections and infestations | 1.640 |
| skin and subcutaneous tissue disorders | 1.562 |
| surgical and medical procedures | 1.226 |
| metabolism and nutrition procedures | 1.018 |
| vascular disorders | 1.016 |
| renal and urinary disorders | 0.890 |
| immune system disorders | 0.884 |
| eye disorders | 0.669 |
| hepatobiliary disorders | 0.603 |
| neoplasms | 0.566 |
| product issues | 0.493 |
| ear and labyrinth | 0.339 |
| blood and lymph | 0.338 |
| congenital, familial, genetic | 0.284 |
| reproductive system and breast | 0.252 |
| pregnancy, puerperium, perinatal | 0.185 |
| endocrine | 0.112 |
